# Sample pooling on triplets to speed up SARS-CoV-2 diagnosis using CDC FDA EUA RT-qPCR kit

**DOI:** 10.1101/2020.06.29.20142836

**Authors:** Byron Freire-Paspuel, Patricio Vega-Mariño, Alberto Velez, Marilyn Cruz, Miguel Angel Garcia-Bereguiain

## Abstract

The CDC designed “FDA Emergeny Use Autorization” 2019-nCoV CDC RT-qPCR kit uses 3 different FAM probes for SARS-CoV-2 diagnosis so 3 reactions per sample are needed. We herein describe a sample pooling protocol: 3 RNA extractions are combined into a single PCR reaction. The sensitivity for this protocol is 100% as no shift on Ct values for N1 or N2 probes were observed. For a typical 96-well plate, triplet assay allows 96 samples processing, speeding up diagnosis.

## Introduction

Several in vitro diagnosis RT-qPCR kits are available on the market for the detection of SARS-CoV-2. Some of them have received emergency use authorization (EUA) from the U.S. Food & Drug Administration (FDA), like 2019-nCoV CDC EUA from the USA Center for Diseases Control and Prevention (CDC). The CDC assay is based on N1 and N2 probes to detect SARS-CoV-2 and RNaseP as an RNA extraction quality control (1). According to CDC protocol for 2019-nCoV CDC EUA, the 3 probes are FAM labelled so 3 PCR reactions are needed for each specimen diagnosis. With no triplex PCR protocol validated for N1, N2 and RNaseP, the current CDC protocol reduces daily sampling processing capacity for a typical 96 well plate PCR device. On developing countries like Ecuador, most of clinical microbiology laboratories running SARS-CoV-2 diagnosis operates with a single Real Time PCR device. Under this scenario, pooling samples while keeping sensitivity is a powerful tool to increase SARS-CoV-2 testing capacity. Also, testing costs are reduced and supply shortage may be mitigated by using a pooling sample protocol, crucial to support surveillance at developing countries.

This study evaluates the performance of a sample pooling RT-qPCR protocol where 3 RNA samples (“triplet”) are loaded into the same RT-qPCR reaction for SARS-CoV-2 diagnosis by using 2019-nCoV CDC EUA kit (IDT, USA).

## Methods

### Study setting

114 clinical specimens (nasopharyngeal swabs collected on 0.5mL TE pH 8 buffer) from individuals selected during SARS-CoV-2 surveillance in Galapagos Islands started on April 8th 2020, were included on the evaluation study. Also, eight negative controls (TE pH 8 buffer) were included as control for carryover contamination. “LabGal” at “Agencia de Regulacion y Control de la Bioseguridad y Cuarentena para Galapagos” at Puerto Ayora in Galapagos Islands (Ecuador) is the only available SARS-CoV-2 diagnosis laboratory on site, operating with a single 96 well plate PCR device (CFX96 from BioRad) to cover a population above 25.000 people.

### RNA Extraction and RT-qPCR for SARS-CoV-2 diagnosis

Samples were tested following an adapted version of the CDC protocol: (1) using PureLink Viral RNA/DNA Mini Kit (Invitrogen, USA) as an alternate RNA extraction method; (2) using CFX96 BioRad instrument. We performed this protocol for 38 SARS-CoV-2 positive and 76 negative samples individually, but also pooling one positive sample with two negative samples at the RT-PCR reaction mix. While 4 uL of a single RNA was added to a single RT-PCR reaction, 2 uL of each RNA was added to the triplet RT-PCR reaction.

### Statistics

For statistical analysis of Ct values, t-student test was performed using Excel.

### Ethics statement

All samples have been submitted for routine patient care and diagnostics. The study was approved by the “Comité de Operaciones Especiales Regional de Galápagos” that is leading board for the Covid19 surveillance in Galapagos Islands. No extra specimens were specifically collected for this validation study. All data used in the current study was anonymized prior to being obtained by the authors.

## Results

We found no significant differences for Ct values between the single and triplet RT-qPCR reaction: 31,30 ± 3,69 vs 31,16 ± 4,04 for N1 (p= 0.72); 34,09 ± 3,83 vs 33,25 ± 3,96 for N2 (p=0.14). Results are detailed on Table 1 and 2. The assay was validated to detect 10 viral RNA copies/uL by using 2019-nCoV N positive control (IDT, USA). All 38 samples that tested positive for the single sample RT-qPCR were also positive for the triplet RT-qPCR, so the sensitivity triplet sample pooling protocol was 100%.

**Table 1.** Average Ct values for N1 and N2 for samples tested on single and triplet RT-qPCR protocol.

|  | N1 |  | N2 |  |
| --- | --- | --- | --- | --- |
|  | Triplet PCR | Single PCR | Triplet PCR | Single PCR |
|  | CT value | CT value | CT value | CT value |
| Mean ± SD | 31,16 ± 4,04 | 31,30 ± 3,69 | 33,25 ± 3,96 | 34,09 ± 3,83 |

**Table 2.** Ct values for N1 and N2 for single and triplet RT-qPCR protocol for the 38 SARS-CoV-2 positive samples included on the study.

| n | ID | N1 Triplet | N1 Single | N2 Triplet | N2 Single |
| --- | --- | --- | --- | --- | --- |
|  |  | PCR | PCR | PCR | PCR |
|  |  | CT value | CT value | CT value | CT value |
| 1 | OCOL | 30,09 | 30,60 | 33,86 | 32,28 |
| 2 | 347 | 32,34 | 31,02 | 36,47 | 32,46 |
| 3 | 351 | 36,84 | 35,87 | 40,56 | 38,72 |
| 4 | 356 | 36,13 | 34,43 | 38,64 | 35,84 |
| 5 | 365 | 33,19 | 32,42 | 36,50 | 35,08 |
| 6 | 783 | 34,74 | 33,57 | 38,37 | 36,03 |
| 7 | 878 | 34,74 | 35,91 | 38,37 | 41,72 |
| 8 | 448 | 30,27 | 29,71 | 33,30 | 31,39 |
| 9 | 449 | 28,26 | 27,15 | 32,21 | 29,33 |
| 10 | 668 | 27,09 | 25,15 | 30,37 | 26,59 |
| 11 | 676 | 27,09 | 36,26 | 30,37 | 39,87 |
| 12 | 683 | 27,09 | 35,45 | 30,37 | 38,08 |
| 13 | 864 | 34,61 | 34,04 | 37,12 | 35,83 |
| 14 | 1102 | 35,32 | 34,05 | 35,51 | 34,43 |
| 15 | I2 | 36,69 | 35,49 | 36,29 | 36,24 |
| 16 | I13 | 37,21 | 36,35 | 36,43 | 36,45 |
| 17 | 983c | 28,01 | 27,01 | 30,90 | 28,19 |
| 18 | 985c | 23,84 | 22,61 | 25,50 | 24,02 |
| 19 | 986c | 34,65 | 33,90 | 36,85 | 37,34 |
| 20 | 988c | 35,73 | 34,76 | 36,18 | 36,80 |
| 21 | 989c | 36,52 | 35,81 | 37,07 | 37,05 |
| 22 | 991c | 30,10 | 28,80 | 30,73 | 29,98 |
| 23 | 997c | 29,76 | 28,27 | 31,19 | 29,82 |
| 24 | 1008c | 29,48 | 28,11 | 31,68 | 29,54 |
| 25 | 1009c | 31,86 | 30,64 | 33,48 | 32,08 |
| 26 | 943 | 30,66 | 32,41 | 32,89 | 36,46 |
| 27 | 958 | 27,77 | 27,45 | 28,61 | 32,80 |
| 28 | 965 | 24,18 | 26,70 | 25,67 | 32,92 |
| 29 | 966 | 33,36 | 32,76 | 35,22 | 37,49 |
| 30 | 967 | 29,89 | 31,34 | 31,87 | 36,15 |
| 31 | 968 | 26,63 | 27,30 | 29,07 | 31,96 |
| 32 | 977 | 31,38 | 30,40 | 34,03 | 35,31 |
| 33 | 992 | 25,57 | 26,06 | 27,12 | 30,08 |
| 34 | 997 | 31,60 | 33,11 | 33,88 | 38,40 |
| 35 | 999 | 27,97 | 29,86 | 29,39 | 34,04 |
| 36 | 1008 | 24,33 | 26,86 | 25,67 | 32,46 |
| 37 | 1009 | 30,70 | 31,90 | 33,00 | 35,00 |
| 38 | 989c(2) | 38,30 | 35,81 | 38,72 | 37,05 |

## Discussion

Our results support the use of a triplet RT-qPCR protocol for SARS-CoV-2 diagnosis without compromising the sensitivity compared to single sample RT-qPCR protocol. This protocol is an easy way to speed up SARS-CoV-2 diagnosis when using the CDC RT-qPCR protocol: the need of three PCR reactions per sample due to FAM labelling for the three probes is corrected by pooling samples in triplets for PCR. This allows to optimize number of samples per running at a typical 96 well PCR device like the one used in our “LabGal” laboratory at Galapagos Islands. For small scale labs at developing countries like Ecuador, this is an alternative way for reagents savings and increase diagnosis capacity without losing sensitivity, and also to compensate supply shortage. Although a few reports regarding sample pooling for SARS-CoV-2 diagnosis have been published on the last weeks (2–6), only 3 of those include sensitivity evaluation (2,3,5). Moreover, this is the first study to our knowledge using CDC FDA EUA RT-qPCR kit and also not showing Ct shifts and reduced sensitivity for sample pooling.

We have been successfully using this protocol during covid19 surveillance at Galapagos Islands where more than 5% (over 1500 subjects) of the population has been tested on a single lab with a single Real Time PRC device within a month period (confirming also a 100% specificity as all positives triplet pools always yielded at least a positive sample). The main limitation of our protocol is the need for running an extra RT-PCR reaction for positives triplets on a single sample mode that delays diagnosis a few hours. So this protocol would not be useful when high prevalence of SARS-CoV-2 is expected and diagnosis is expedited as for hospitalized individuals. However, when a low prevalence is expected and a wide screening is the goal, the triplet protocol would be of great help.

## Data Availability

All relevant data is included on the manuscript

## Acknowledgments

We thank the medical personnel from “Ministerio de Salud Pública” at Galapagos Islands and the staff from the “Agencia de Regulación y Control de la Bioseguridad y Cuarentena para Galápagos” for their support. We specially thank Gabriel Iturralde, Oscar Espinosa and Dr Tannya Lozada from “Dirección General de Investigación de la Universidad de Las Américas”, and the authorities from Universidad de Las Américas, for logistic support to make SARS-CoV-2 diagnosis possible in Galapagos Islands.

